## Extended Data Figures for "Head-to-head comparison of aptamer- and antibody-based proteomic platforms in human cerebrospinal fluid samples from a real-world memory clinic cohort"

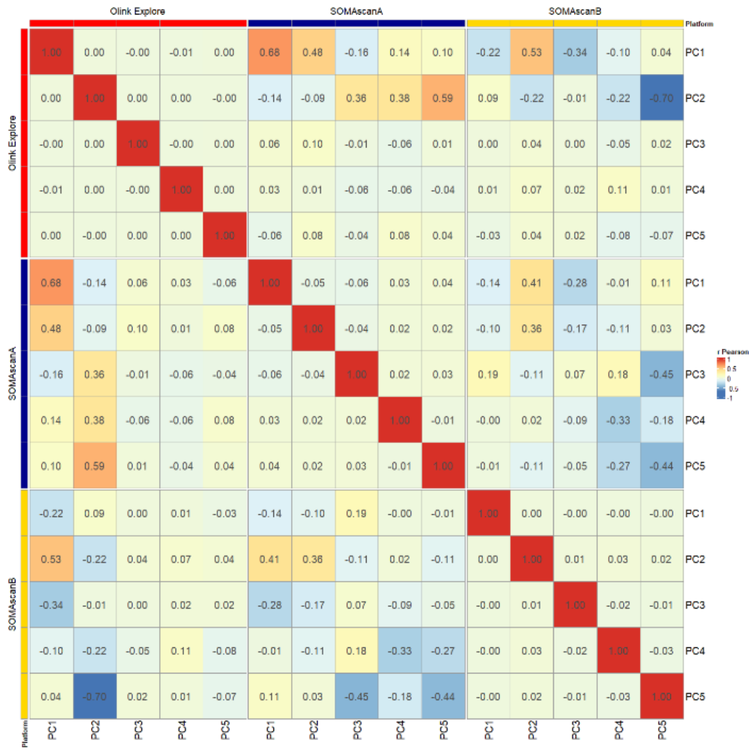


***Supplementary Figure 1. Pearson correlation between PCs across proteomic analysis in Olink Explore and SOMAscan.***

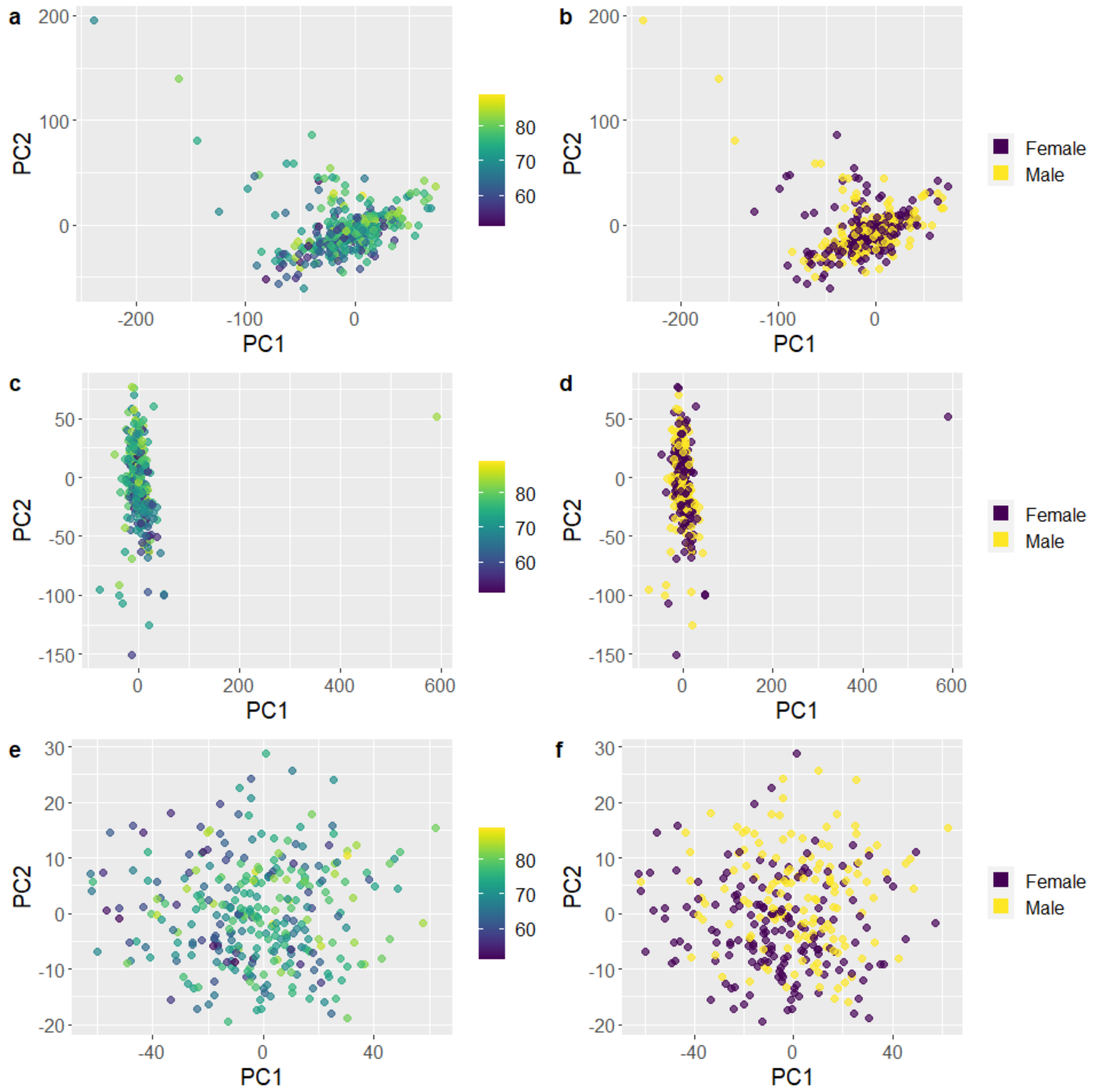


***Supplementary Figure 2. Representation of PC1 and PC2 coloured by Age at LP and sex on SOMAscan and Olink datasets**. SOMAscanA datasets (a and b), SOMAscanB datasets (c and d) and Olink Explore datasets (e and f). Outlier individuals were not considered in this plot.*

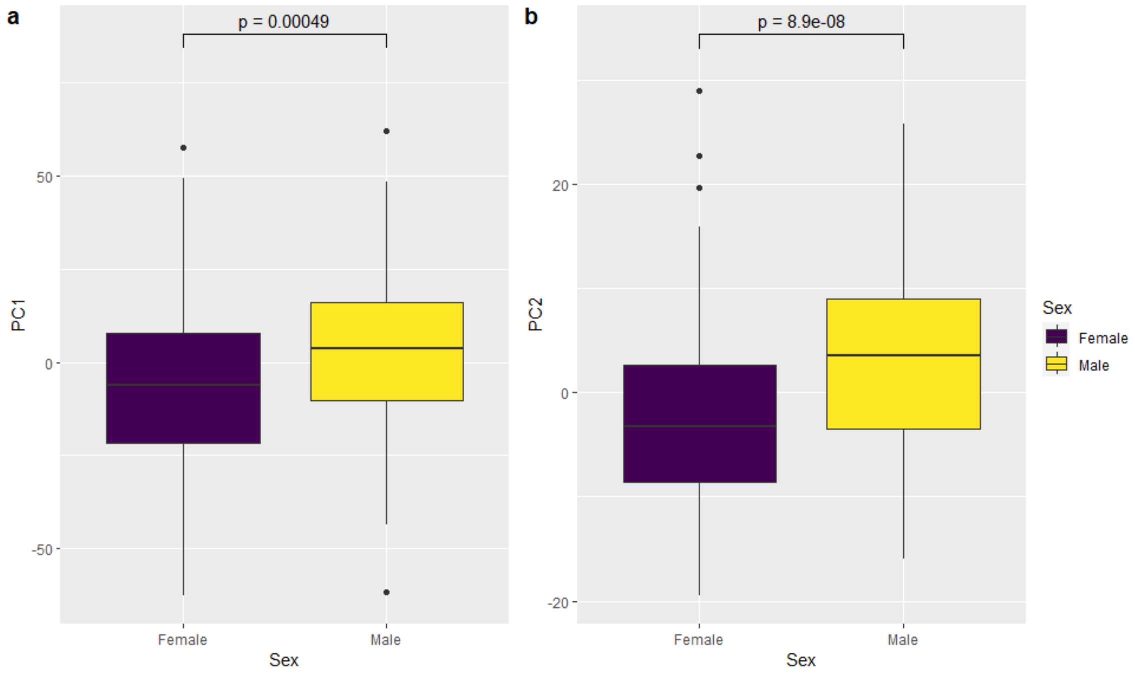


***Supplementary Figure 3. Differences in PC levels between sex in the Olink Explore proteomic platform***. *left: PC1. right: PC2.*

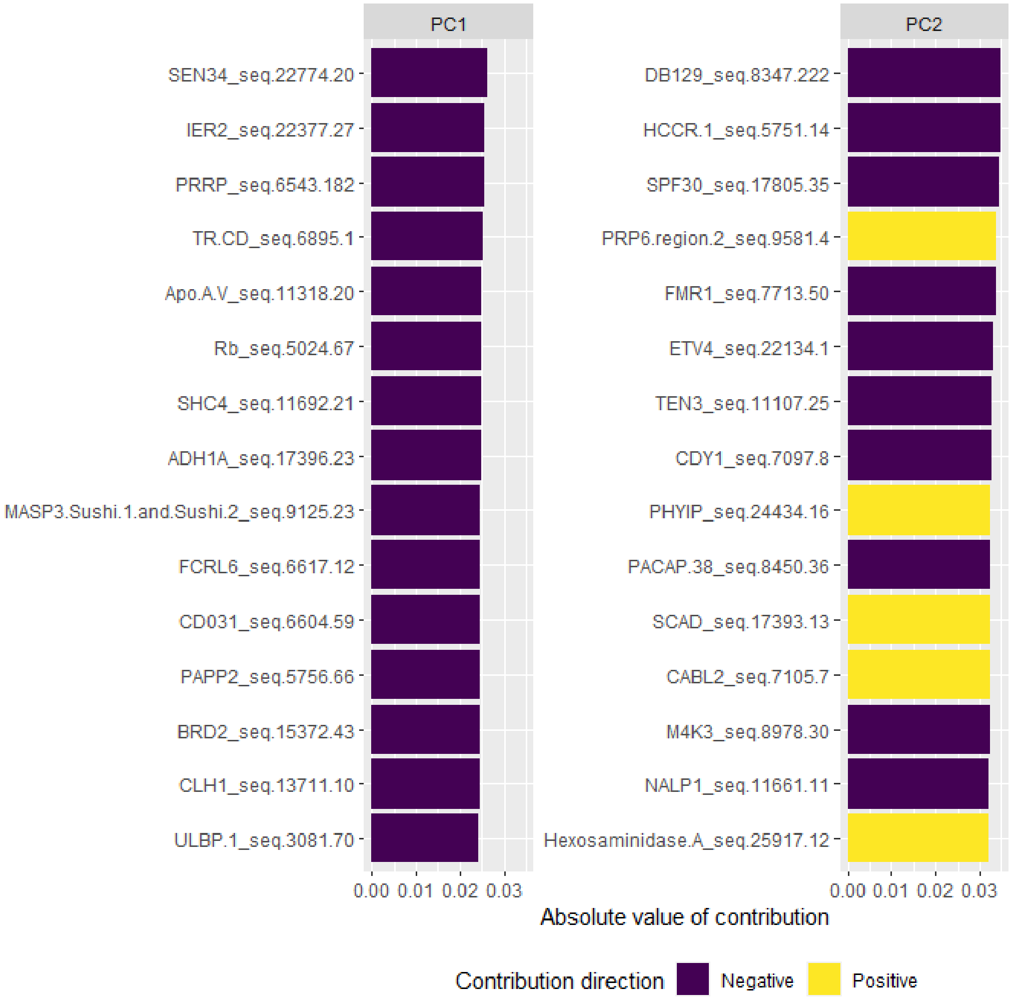


***Supplementary Figure 4. Top 15 proteins contributing to PC1 and PC2 in the SOMAscanA dataset.***

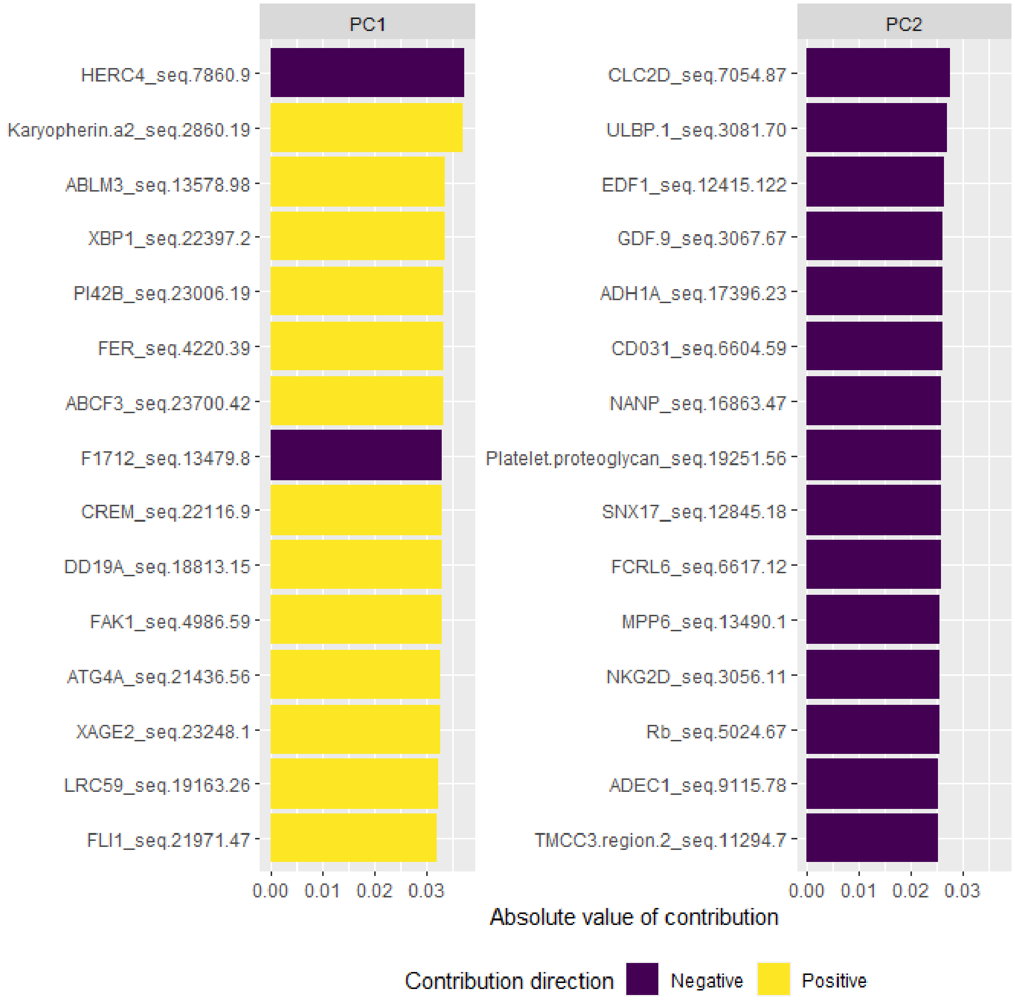


***Supplementary Figure 5. Top 15 proteins contributing to PC1 and PC2 in the SOMAscanB dataset.***

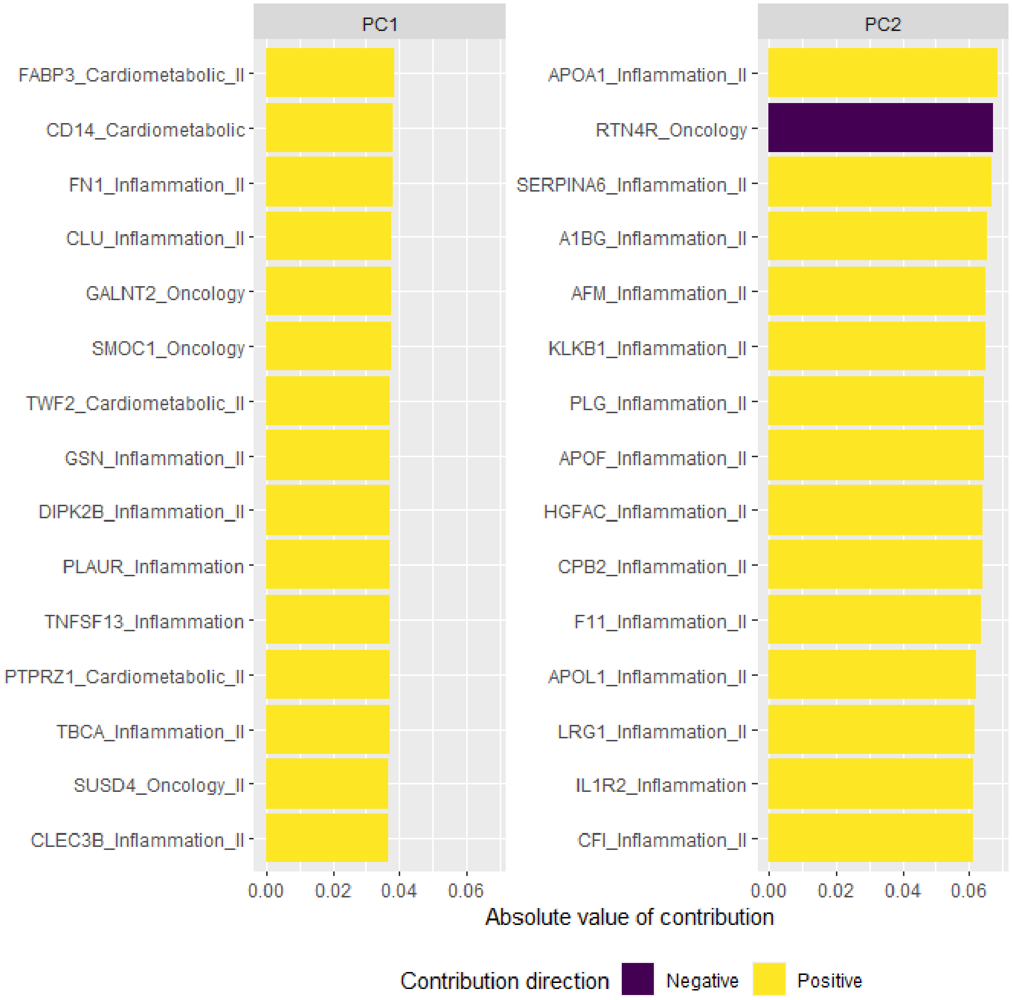


***Supplementary Figure 6. Top 15 proteins contributing to PC1 and PC2 in the Olink Explore dataset.***

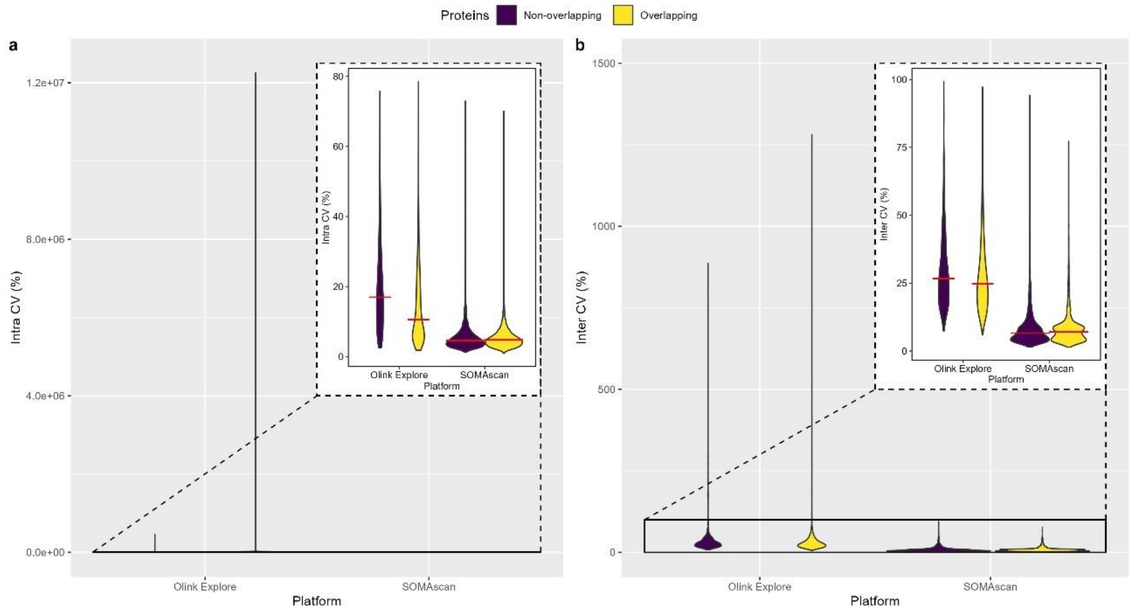


***Supplementary Figure 7. Coefficient of variation for SOMAscan and Olink Explore Platforms****. a) Intra- and b) Inter-assay coefficient of variation coloured by the overlapping between SOMAscan and Olink assays. The red line in the zoom plot represents the median CV for each platform.*

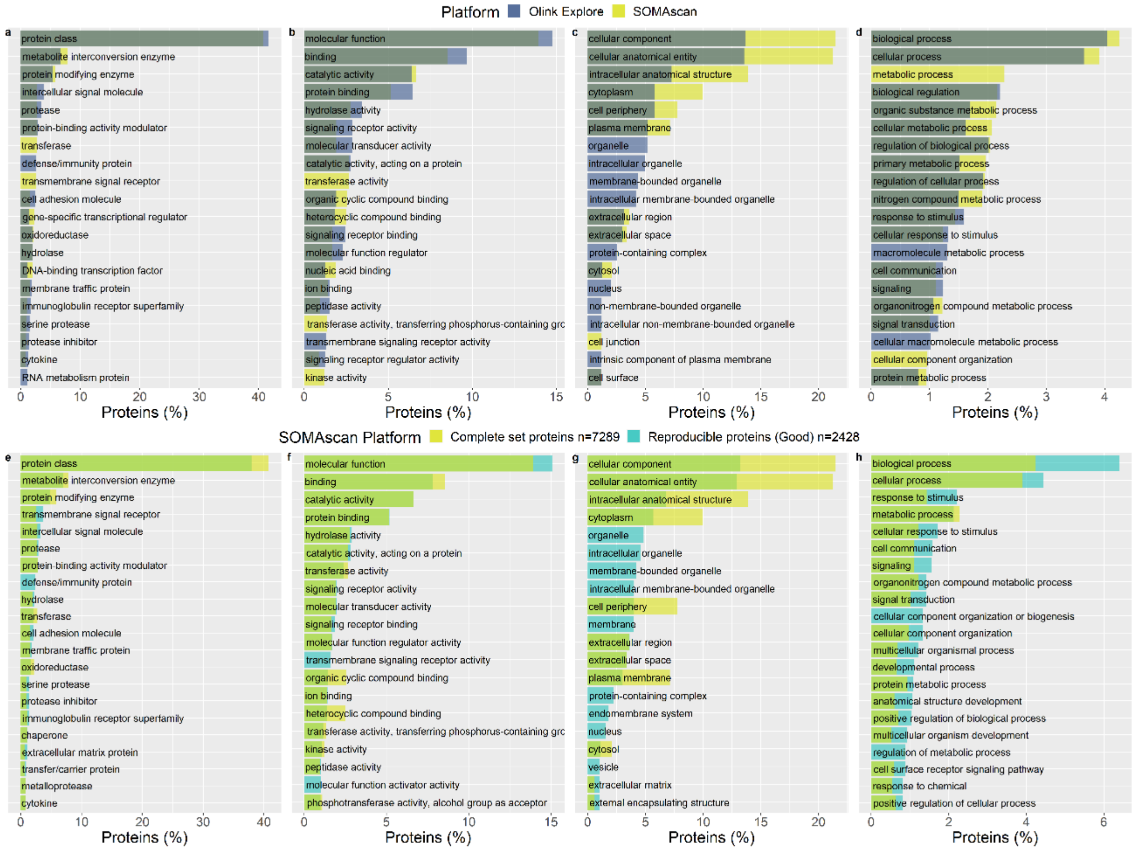


***Supplementary Figure 8*. *Top 20 significant classifications in PANTHER database (FDR < 0.05).*** *A) protein class, B) molecular function, C) cellular compartment and D) biological process in Olink® Explore (dark blue; n=2,872) and SOMAscan® (yellow; n=6,218). E) protein class, F) molecular function, G) cellular compartment and H) biological process in the complete set of SOMAscan® proteins (yellow; n=6,218) and reproducible SOMAscan® proteins with Good intra-assay correlation (rho ≥ 0.5) (light blue; n=2,120).*

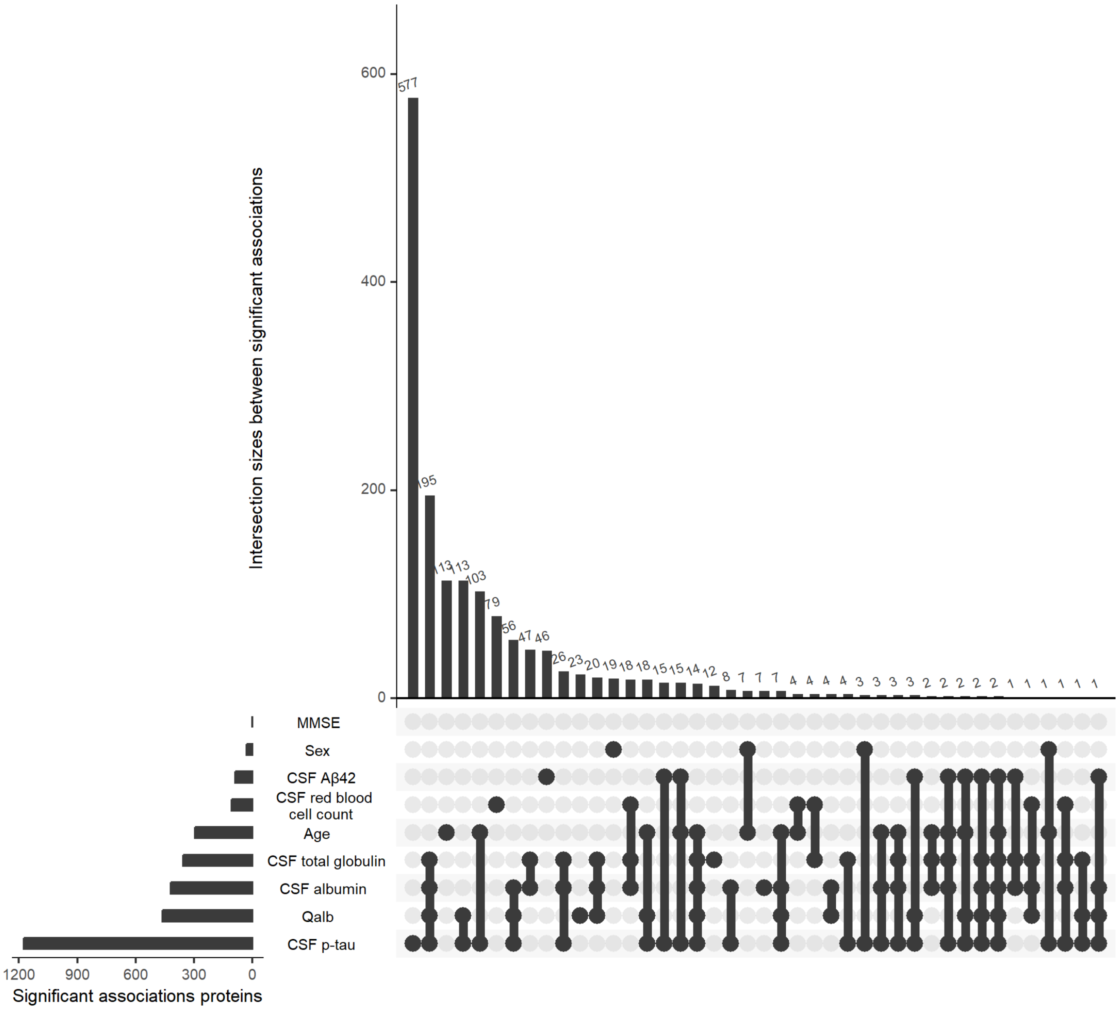


***Supplementary Figure 9. Representation of the overlapping significant proteins among the association analyses with CSF Biological Traits, Sample Demographics, and Alzheimer's Disease Endophenotypes****. This graph was conducted using the UpSetR R package.*

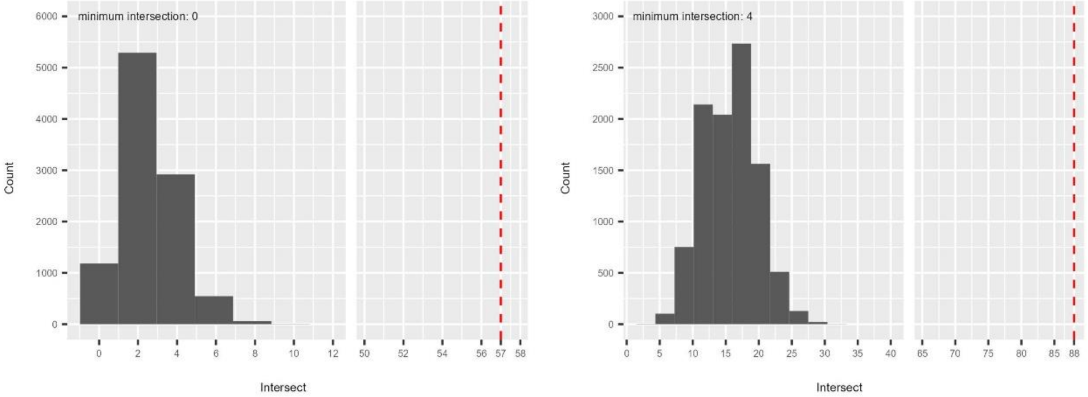


***Supplementary Figure 10. Simulation of the overlapping between three sets of proteins.*** *Left) Considering the complete set of SOMAscan proteins n=7,289. Right) Considering only reliable and reproducible proteins with Good intra-assay correlation n=2,428. The x-axis is truncated to provide a better visualization of results. The red dashed line represents the number of overlapping proteins that we found in our analysis.*

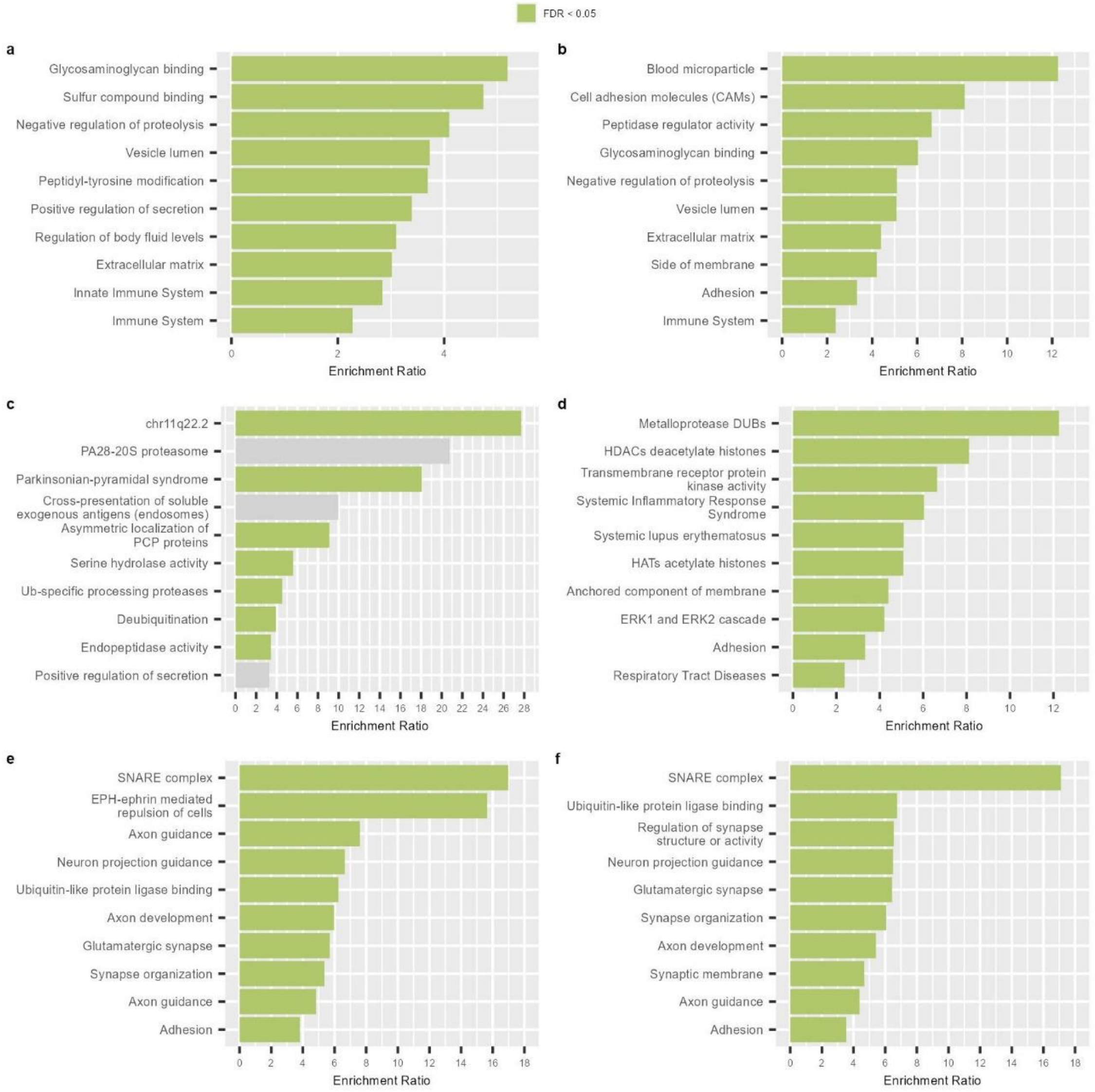


***Supplementary Figure 11. Top 10 enrichment analysis of non-intersecting proteins in the top 500 ranking of significant associations of MMSE, CSF Aβ42 and CSF p-tau.*** *A) 332 non-intersecting proteins in the MMSE top ranking considering the complete set of proteins, B) 264 non-intersecting proteins in the MMSE top ranking considering reproducible SOMAscan proteins, C) 240 non-intersecting proteins in the CSF Aβ42 top ranking considering the complete set of proteins and D) 190 non-intersecting proteins in the CSF Aβ42 top ranking considering reproducible SOMAscan proteins, E) 315 non-intersecting proteins in the CSF p-tau top ranking considering the complete set of protein, and F) 260 non-intersecting proteins in the CSF p-tau top ranking considering reproducible SOMAscan proteins. We used the WebGestalt tool for the enrichment analysis.*

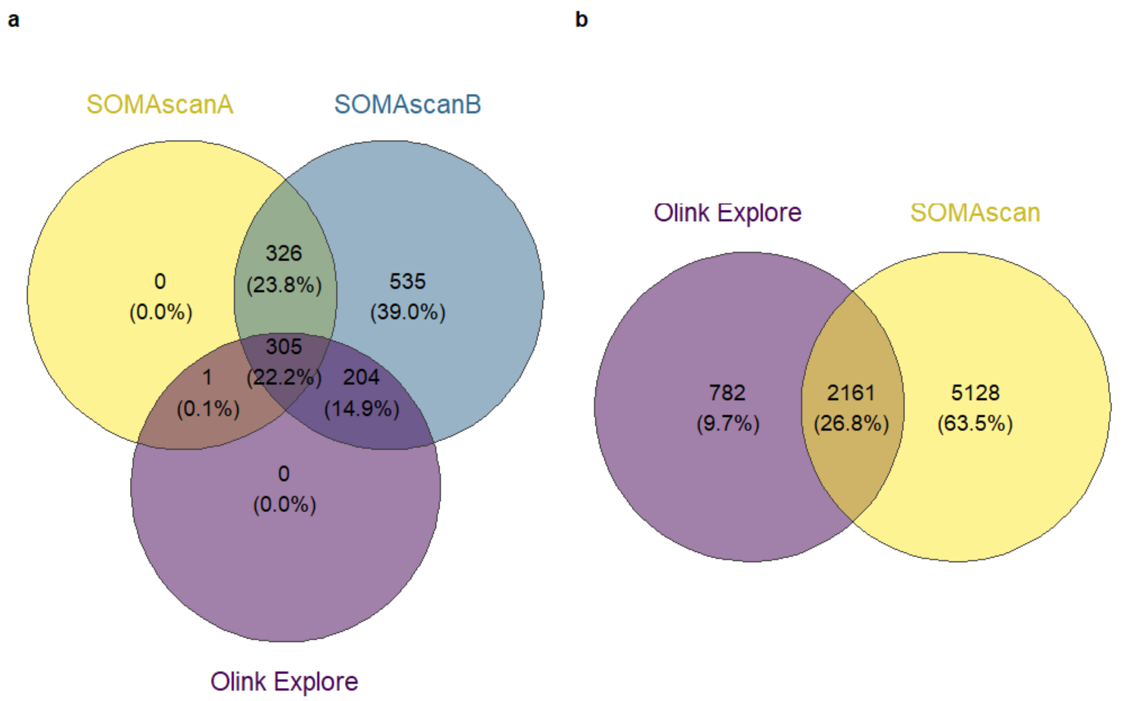


***Supplementary Figure 12. Venn diagrams.*** *A) Individuals overlapping in SOMAscanA, SOMAscanB and Olink Explore proteomic analyses. B) Proteins identified in SOMAscan and Olink Explore platforms, we performed the pairing of both platforms by UniProtIDs.*
